# Pervasive Downward Bias in Estimates of Liability Scale Heritability in GWAS Meta-Analysis: A Simple Solution

**DOI:** 10.1101/2021.09.22.21263909

**Authors:** Andrew D. Grotzinger, Javier de la Fuente, Michel G. Nivard, Elliot M. Tucker-Drob

## Abstract

SNP heritability is a fundamental quantity in the genetic analysis of complex traits. For binary phenotypes, in which the continuous distribution of risk in the population is unobserved, observed-scale heritabilities must be transformed to the more interpretable liability-scale. We demonstrate here that the field standard approach for performing the liability conversion can downwardly bias estimates by as much as ∼20% in simulation and ∼30% in real data. These attenuated estimates stem from the standard approach failing to appropriately account for varying levels of ascertainment across the cohorts comprising the meta-analysis. We formally derive a simple procedure for incorporating cohort-specific ascertainment based on the summation of effective sample sizes across the contributing cohorts, and confirm via simulation that it produces unbiased estimates of liability-scale heritability.

---

*SNP heritability* 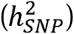 quantifies the proportion of total variance in a phenotype within a population that is attributable to the additive effect of tagged genetic variants. For continuously measured quantitative traits, in which phenotypic variation is directly observed, 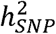 estimates produced from standard methods such as LD score regression (LDSC)^1^ are directly interpretable. However, when the measured phenotypes are binary (e.g., for case-control disease traits) conventional estimates of 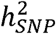 are not easily interpreted for two reasons. The first is due to the binarized scale of the data, where 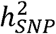 is most interpretable when taking into account the continuous distribution of risk in the population. The second relates to the fact that GWAS of disease traits are often performed on ascertained samples, in which affected individuals are overrepresented so as to increase statistical power for rare disorders. The standard transformation for binary traits then uses a liability threshold model to convert observed-scale SNP heritability 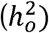 to liability-scale SNP heritability 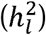 in order to produce an estimate that both accounts for the continuous distribution of risk in the population and is not biased by ascertainment. In practice, 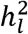 is commonly estimated with summary-based methods such as LDSC using results from GWAS meta-analysis across many different samples, varying in their levels of ascertainment.

Here we highlight a critical error in the standard practice for calculating 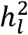 from GWAS meta-analysis that can cause substantial downward bias due to variation in cohort-specific ascertainment, and we formally derive a simple procedure for obtaining unbiased 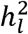 estimates. We report results from simulations that illustrate the extent of the downward bias across a variety of conditions, and showcase the unbiased nature of the proposed procedure within these same conditions. We go on to quantify the extent of this bias in 12 recent GWAS meta-analyses of case/control psychiatric and neurological traits. It appears that the biased approach has been used for 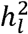 estimates for nearly all meta-analytic GWAS of binary traits, with the exception of Mullins et al. (2021)^2^ who implemented an unbiased estimator of 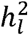 akin to the procedure developed here. We report the corrected estimates of 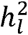 under the same population prevalences assumed by the original GWAS reports and we observe that the field standard approach for computing 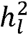 attenuates estimates by as much as 31%.

Observed scale heritability is estimated within univariate LDSC as:^1^

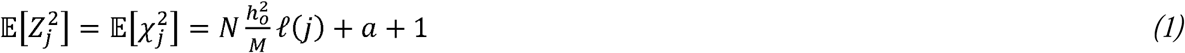

where *N* is the sample size, 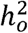 is the observed scale heritability, ℓ(*j*) is the total LD-score for SNP *j, M* is the total number of SNPs used to calculate the LD-scores, and *a* is a term representing unmeasured sources of confounding such as population stratification.

When summary data are derived from a single case-control GWAS (either of a single sample or of raw data that have been combined across multiple samples prior to GWAS), the observed scale heritability 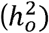 can be converted to the liability scale 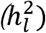 as follows:^3–5^

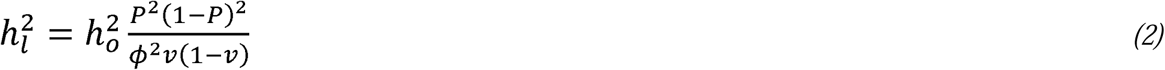

where *ν* is the sample prevalence, *P* is the population prevalence, and *ϕ* is the height of the standard normal probability density function at the threshold corresponding to *P*. Combining *Eqs. 1* and *2* produces the reduced form LDSC equation for binary traits:

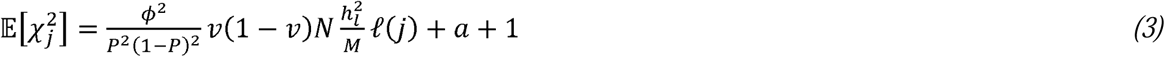

In the **Supplementary Note**, we show that when GWAS summary data are derived from meta-analysis of summary statistics from multiple, individual case-control GWAS, the appropriate reduced form equation for estimating 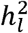 is:

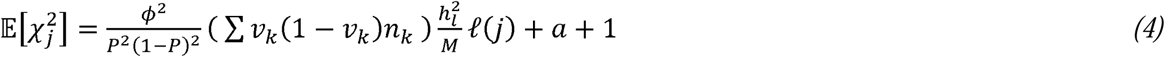

which resembles *Eq. 3* for summary statistics derived from a single GWAS, with the key difference being that *ν* (1 − *ν*)*N* is replaced by (∑*ν*_*k*_(1 − *ν*_*k*_)*n*_*k*_).

Importantly, currently available software does not allow for direct entry of (∑*ν*_*k*_(1 − *ν*_*k*_)*n*_*k*_), and standard practice in LDSC analysis of meta-analytic GWAS summary data has been to compute a single meta-analytic *ν* as the total sample prevalence (i.e. aggregate number of cases across all samples divided by the aggregate sample size, and enter this quantity into *Eq. 3*). When samples are differentially ascertained, as is nearly always the case in empirical settings, such an approach is not equivalent to the correct approach given by *Eq. 4*, i.e.:

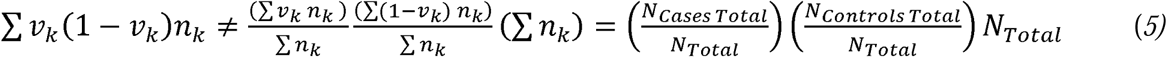

We refer to the quantity on the left of the inequality as the summation of cohort-specific ascertainments, and the quantities on the right of the inequality as the total sample ascertainment. In the **Supplemental Note** we describe a simple procedure in which the correct estimate of 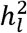 (i.e. *Eq. 3*) can be obtained for meta-analytic summary statistics using standard software (e.g., LDSC,^1^ Genomic SEM,^6^ MTAG^7^ LDAK^8^). First, the effective sample size, *EffN* is computed for each study, *k*, as:

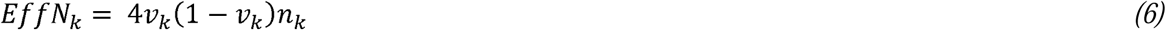

where *EffN* represents the sample size for an equivalently powered GWAS within a balanced sample (i.e. 50% cases, 50% controls). Because the *EffN* values are directly comparable across GWAS samples they can be summed. The sum of *EffN* across all contributing GWAS (∑ *EffN*) is then entered for *N* along with the *ν*=.5, so as to represent the balanced nature of the design. The population prevalence from collateral epidemiological data is entered for *P* as usual.

In addition to being pragmatically appealing given currently available software, this approach has two additional advantages. First, a number of statistical pipelines adopted by major, genomic research consortia (e.g., the RICOPILI pipeline implemented by the Psychiatric Genomics Consortium [PGC]^1^) default to outputting ∑ *EffN* in the GWAS summary statistics output. Second, this allows the user to account for both cohort-specific and SNP-specific information. That is, when participant sample size varies by SNP, as is often the case given different genotyping platforms utilized by cohorts, it is preferrable to use this SNP-specific information.

We used simulation to empirically evaluate the extent of potential bias associated with the field standard, total ascertainment approach, and to confirm the unbiased nature of the cohort-specific ascertainment approach. For all simulations, the liability scale heritability 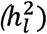 was 15% in the generating population, the population prevalence (*P*) was 1%, and the meta-analytic GWAS consisted of ten cohorts, each with a sample size of 5,000 (i.e., total meta-analytic *N* = 50,000). All cohorts were specified to genetically correlate at 1 to reflect meta-analysis of the same phenotype. Each cohort was also specified to consist of either 50% cases and 50% controls (representing samples with high ascertainment) or 10% cases and 90% controls (representing samples with relatively low ascertainment). These parameters were chosen to reflect a combination of realistic scenarios generally observed for univariate GWAS of binary traits.

We conducted simulations for 11 conditions that iteratively changed with respect to the number of contributing cohorts with 50% cases and controls. More specifically, Condition 1 consisted of 10 cohorts with 10% cases and 90% controls, Condition 2 consisted of 9 cohorts with 10% cases and 90% controls and 1 cohort with 50% cases and controls, and so on up through Condition 11 consisting of 10 cohorts with 50% cases and controls. For each of the 11 conditions, we ran 100 simulations, each comprised of 10 cohort-level sets of summary statistics (i.e., 11,000 simulated cohorts). Summary statistics were generated directly from the LD-score regression equation, subsequently combined across the 10 cohorts using inverse variance weighted (IVW) meta-analysis, and finally analyzed in LDSC in one of two ways (see **Method** for details). The first was using the standard procedure of inputting the total sample prevalence (*v*_*Total*_) and total sample size (*N*_*Total*_). The second was using ∑ *EffN* across the cohorts and a *ν* of .5, as we propose here.

Simulation results are presented in Figure 1 and Table 1. These results reveal three primary findings. First, the field standard approach of using *v*_*Total*_ can produce substantial, downward bias in liability-scale heritability estimates, with bias increasing as a function of the degree of variability in ascertainment across contributing cohorts (Figure 1A). Thus, bias was greatest for those conditions where there was an even mixture of 50%/50% and 10%/90%, case/control cohorts; in these instances, the downward bias was ∼20% (Table 1). Second, both approaches produce the same estimates when ascertainment is equivalent across all cohorts (Figure 1B and 1L). Importantly, the standard procedure of using total sample prevalence (*v*_*Total*_) is not biased as a function of the overall degree of ascertainment. Rather, the bias is attributable to the level of ascertainment differences across cohorts. Third, our proposed procedure of using ∑ *EffN* removes this bias, producing accurate estimates of the population-level, liability scale heritability. Having established that ∑ *EffN* produces an accurate estimate of 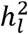, we went on to examine the difference across using *v*_*Total*_ and ∑ *EffN* in real data.

**Table 1.** Simulation Results across Conditions

| *Condition* | **50% Cases /**<br>**50% Controls** | **10% Cases /**<br>**90% Controls** | **Proposed Approach: $\sum EffN$** |  |  | **Field Standard Approach: $v_{Total}$** |  |  |
| --- | --- | --- | --- | --- | --- | --- | --- | --- |
|  |  |  | *Mean $h^2$ (SD)* | *$h^2$ Range* | *Mean % Bias (SD)* | *Mean $h^2$ (SD)* | *$h^2$ Range* | *Mean % Bias (SD)* |
| *Condition 1* | 0 | 10 | 14.83 (0.56) | 13.31 : 16.33 | -1.18 (3.74) | 14.83 (0.56) | 13.31 : 16.33 | -1.18 (3.74) |
| *Condition 2* | 1 | 9 | 14.87 (0.55) | 13.78 : 16.09 | -0.87 (3.65) | 13.09 (0.48) | 12.13 : 14.17 | -12.72 (3.22) |
| *Condition 3* | 2 | 8 | 14.88 (0.49) | 13.76 : 16.37 | -0.78 (3.25) | 12.30 (0.40) | 11.37 : 13.53 | -17.99 (2.69) |
| *Condition 4* | 3 | 7 | 14.87 (0.55) | 11.02 : 15.95 | -0.87 (3.68) | 11.96 (0.44) | 8.87 : 12.83 | -20.27 (2.96) |
| *Condition 5* | 4 | 6 | 14.86 (0.40) | 14.09 : 16.02 | -0.95 (2.67) | 11.89 (0.32) | 11.27 : 12.83 | -20.71 (2.13) |
| *Condition 6* | 5 | 5 | 14.82 (0.33) | 13.91 : 15.77 | -1.19 (2.22) | 12.00 (0.27) | 11.26 : 12.76 | -20.01 (1.80) |
| *Condition 7* | 6 | 4 | 14.87 (0.33) | 14.27 : 15.62 | -0.89 (2.22) | 12.32 (0.28) | 11.83 : 12.95 | -17.85 (1.85) |
| *Condition 8* | 7 | 3 | 14.89 (0.29) | 14.03 : 15.68 | -0.74 (1.93) | 12.77 (0.25) | 12.03 : 13.44 | -14.90 (1.66) |
| *Condition 9* | 8 | 2 | 14.97 (0.29) | 14.26 : 15.93 | -0.23 (1.99) | 13.39 (0.27) | 12.76 : 14.26 | -10.71 (1.78) |
| *Condition 10* | 9 | 1 | 14.86 (0.28) | 13.99 : 15.38 | -1.09 (1.89) | 13.97 (0.27) | 13.18 : 14.49 | -6.82 (1.78) |
| *Condition 11* | 10 | 0 | 14.84 (0.27) | 14.25 : 15.41 | -1.05 (1.78) | 14.84 (0.27) | 14.25 : 15.41 | -1.05 (1.78) |

*Note.* The **50% cases / 50% controls** and **10% cases / 90% controls** columns denote the total number of cohorts with this case/control split for each condition. The **Proposed Approach** columns denote the simulation results when using $\sum EffN$ for the liability correction. The **Field Standard Approach** columns report simulation results when using $v_{Total}$ .

**Figure 1.**
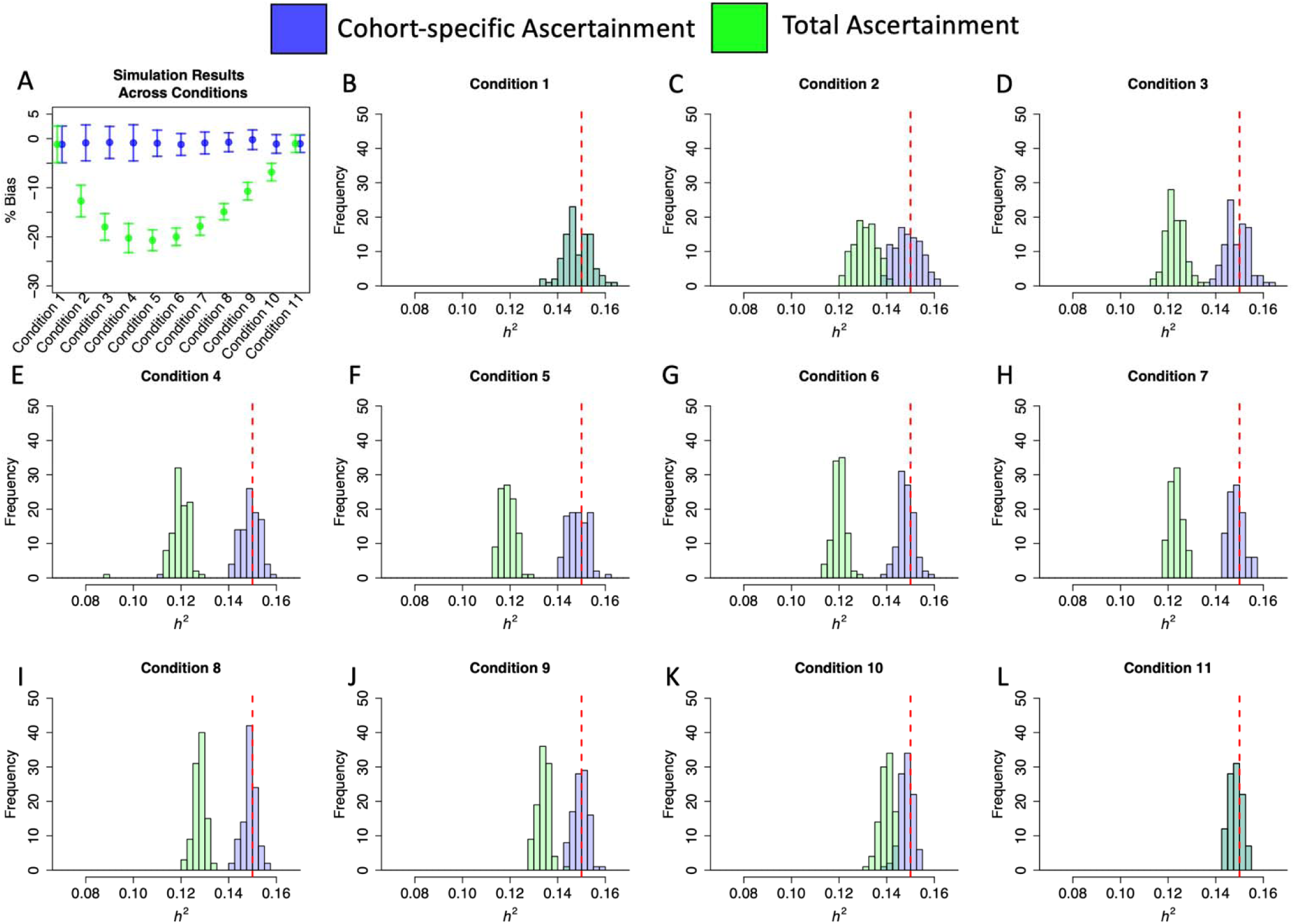
Simulation Results across Conditions. *Panel A* depicts the mean percentage bias on the y-axis across the 11 simulation conditions on the x-axis. Error bars depict +/- 1 *SD. Panels B*-*L* depict the individual point estimates from the 100 simulations per condition across the 11 conditions. The red dashed line indicates the liability scale *h*^*2*^ in the population of 15%. All panels depict in green the results from using to account for cohort-specific ascertainment and in blue the results from using *v*_*Total*_.

We compared the field standard procedure of using *v*_*Total*_ versus our proposed approach of using ∑ *EffN* for 12 major, binary traits for which sufficient cohort-level information was available We used the same population prevalences from the original GWAS publications from which the summary statistics were derived. We quantify bias here as the proportional difference across ∑ *EffN* and *ν*_*Total*_ (*i. e*. .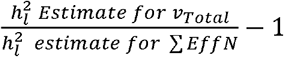). Consistent with simulation findings, real data results revealed that in all cases using *v*_*Total*_ produced a deflated estimate of liability scale heritability relative to ∑ *EffN* This bias ranged from as little as 1.3% for autism spectrum disorder^9^ to as much as 28.1% for alcohol use disorder^10^ and 31.8% for bipolar disorder^2^ (Table 2). In all but one instance, the heritability estimates reported in the corresponding manuscripts most closely matched those produced from using *v*_*Total*_ (Supplementary Table 1). The exception was the most recent release (Freeze 3) of the PGC bipolar summary statistics^2^ which report a liability scale heritability consistent with using ∑ *EffN*.

**Table 2.** LDSC Heritability Estimates Using Total or Cohort-specific Ascertainment

| Trait | Reference | Pop. Prev. | Cases | Controls | Field Standard Approach ($v_{Total}$): $h^2$ (SE) | Proposed Approach ($\sum EffN$): $h^2$ (SE) | % Bias |
| --- | --- | --- | --- | --- | --- | --- | --- |
| ADHD | Demontis *et al.* (2019)<sup>20</sup> | 5.0% | 19,099 | 34,194 | 22.1% (1.4) | 23.7% (1.6) | -6.7% |
| ALCH | Walters *et al.* (2018)<sup>10</sup> | 15.9% | 10,206 | 28,480 | 10.0% (1.8) | 13.9% (2.5) | -28.1% |
| ALZ | Kunkle *et al.* (2019)<sup>21</sup> | 4.3% | 21,982 | 41,944 | 5.8% (0.9) | 7.0% (1.1) | -20.7% |
| AN | Watson *et al.* (2019)<sup>22</sup> | 0.9% | 16,992 | 55,525 | 13.8% (0.9) | 15.5% (1.0) | -10.8% |
| ASD | Grove *et al.* (2019)<sup>9</sup> | 1.2% | 18,381 | 27,969 | 11.7% (1.0) | 11.9% (1.0) | -1.3% |
| BIP | Mullins *et al.* (2021)<sup>2</sup> | 2.0% | 41,917 | 371,549 | 12.8% (0.5) | 18.7% (0.8) | -31.8% |
| CUD | Johnson *et al.* (2020)<sup>23</sup> | 1.0% | 14,080 | 343,726 | 6.7% (0.6) | 7.5% (0.7) | -11.5% |
| MDD | Wray *et al.* (2018)<sup>24</sup> | 15.0% | 59,851 | 113,154 | 10.2% (0.6) | 11.5% (0.7) | -11.8% |
| OCD | Arnold *et al.* (2018)<sup>25</sup> | 2.5% | 2,688 | 7,037 | 28.5% (4.4) | 29.9% (4.6) | -4.7% |
| PTSD | Nievergelt *et al.* (2019)<sup>26</sup> | 30.0% | 23,212 | 151,447 | 5.3% (0.9) | 6.1% (1.1) | -12.9% |
| SCZ | Ripke *et al.* (2020)<sup>27</sup> | 1.0% | 53,386 | 77,258 | 20.7% (0.7) | 22.3% (0.8) | -6.9% |
| TS | Yu *et al.* (2019)<sup>28</sup> | 0.8% | 4,819 | 9,488 | 21.5% (2.5) | 22.4% (2.6) | -4.0% |

SNP heritability is a fundamental quantity in complex trait genetics. As such, SNP heritability estimates from GWAS summary statistics are standard results to report in any major GWAS meta-analysis effort. For binary traits, such as case-control disease traits, SNP heritability estimates must be converted to the liability scale in order to be meaningfully interpreted. We demonstrate here that the de facto field standard approach for estimating liability scale heritability from meta-analytic GWAS summary data can downwardly bias these estimates by as much as ∼20% in simulation and ∼30% in real data. We have introduced a simple procedure for obtained unbiased estimates of liability scale SNP heritability in these contexts.

Downwardly biased estimates of SNP heritability will propagate to produce downwardly biased estimates of genetic covariance, which may in turn bias methods that rely on these estimates (e.g., MTAG,^11^ Genomic SEM^6^). Importantly, genetic correlations are expected to be unaffected by this bias as they standardize genetic covariance estimates relative to heritability estimates, thereby cancelling out the bias. Another issue which merits further investigation is the presence of ascertainment differences that stratify by meaningful covariates across cohorts. For example, it is currently unknown how estimates may be biased when ascertainment varies across GWAS cohorts more for one sex than the other.

Genomic-relatedness matrix restricted maximum likelihood (GREML)^3,12^ is a major alternative to LDSC that estimates heritability using raw genotypes among unrelated individuals. While LDSC has the advantage of requiring only summary-level data, and is thus especially applicable to GWAS meta-analysis, GREML is often considered preferable when raw data are available,^13,14^ as it is often found to produce larger SNP heritability estimates than those obtained from LDSC.^15^ One potential explanation for this discrepancy includes the possibility that, because LDSC is typically applied to meta-analytic GWAS data, it will only detect the portion of heritable signal that is consistent across contributing GWAS datasets. A second potential explanation for this discrepancy is that LDSC may produce attenuated heritability estimates due to discrepancies between LD structure in the reference data used to construct the LD scores and the samples from which the GWAS estimates were derived. While these issues are still likely to be at play, the present findings highlight another source of discrepancy across LDSC and GREML for binary traits that is easily corrected.

The discrepancy between a heritability estimate from GWAS data and those reported from family-based (e.g., twin) studies is referred to as missing heritability. Missing heritability is in and of itself often used as a meaningful metric to the extent that it provides a measure of the genetic signal left to uncover, for example, in the rare variant end of the allelic spectrum.^16^ Liability scale heritability estimates that are downwardly biased to the degree observed here will likely shift the level of missing heritability for binary outcomes. Indeed, current reports of SNP-based heritability recapturing only 30%-50% of the heritability estimates from twin studies of psychiatric disorders (i.e., 70%-50% missing heritability)^17^ may need to be revisited for binary traits from which heritability estimates are derived from GWAS meta-analysis. To this end, we strongly recommend using ∑ *EffN* for liability correction, as we have shown this to produce an unbiased estimate of SNP-based heritability in the population.

## Method

### Simulation of SNP Heritability

#### Simulation of Summary Statistics

Each simulation began by generating genome-wide summary statistics for binary traits for 10 individual cohorts. All simulations specified a population prevalence of 1%, a liability scale heritability of 15% in the population, cross-trait intercepts of 0 to reflect no sample overlap across the 10 cohorts, and a univariate intercept of 1.04 to reflect minimal, uncontrolled for population stratification. Each cohort was specified to have a sample prevalence of either 10% (low ascertainment) or 50% (high ascertainment), with the balance of cohorts with low and high ascertainment varying across 11 simulation conditions (see Table 1 for details on each condition). For each condition, 100 sets of 10 cohorts were simulated (i.e., 1,000 cohort level summary statistics per condition for a total of 11,000 simulated cohorts across the 11 conditions). Note that when liability scale heritability is equal, but sample prevalence differs across cohorts, observed scale heritability will differ across cohorts. Data were simulated using European population LD scores provided by the original LDSC developers^18^ for 1,184,461 HapMap3 SNPs, excluding the MHC region, according to simulation procedures first described in de la Fuente et al. (2021).^19^ More specifically, summary statistics were simulated following the multivariate LDSC equation:

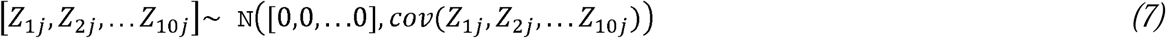

where

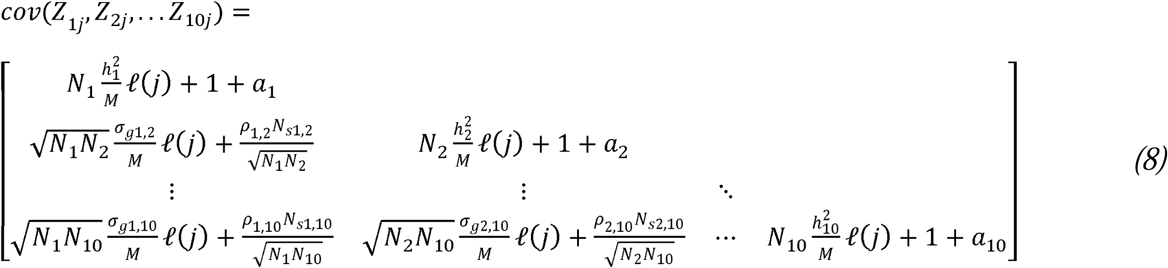

and [*Z*_1*j*_, *Z*_2*j*_, … *Z*_10*j*_]reflects the *Z* statistics for the 10 GWAS cohorts (expressed in condensed form, not depicting cohorts 3-9 from the current simulations for display reasons), *M* is the number of SNPs from the LD file (1,184,461), *N*_*s*_ is the number of overlapping individuals (0), *N* is the sample size of the individual GWAS (5,000), ρ is the phenotypic correlation (0.5), *ℓ(j)* is the LD score of SNP *j*, and *a+*1 reflects the univariate LDSC intercept that picks up on unmeasured confounds, such as population stratification (1.04). The bivariate LDSC intercept, expressed as 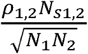 for cohorts 1 and 2, was 0 owing to setting the sample overlap (*N*_*s*_) to 0 for all simulations. GWAS *Z*-statistics were simulated following the equation above and utilizing the *mvrnorm* R function from the *MASS* package for each SNP.

From the simulated cohort-level GWAS *Z*-statistics, we computed logistic betas as follows:

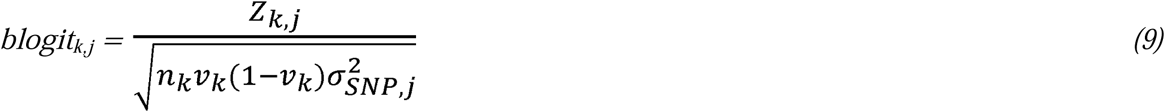

where *v*_*k*_ and *n*_*k*_ reflects the cohort-specific sample prevalence and sample size, respectively, and 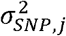 reflects the variance of a given SNP *j* calculated as *2*MAF*(1-MAF)*, where *MAF* is the minor allele frequency. The logistic standard errors for a given SNP *j* and cohort *k* were then calculated as:

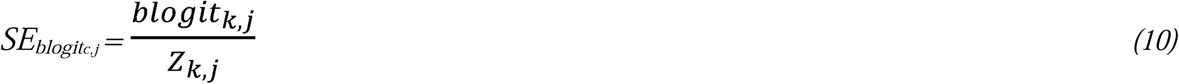

These logistic betas and standard errors were used to calculate the inverse-variance weighted (IVW) meta-analytic beta across the 10 contributing cohorts as described in the **Supplementary Note**. This procedure then produced a single summary statistics file reflecting the meta-analyzed output across the 10 simulated cohorts. This summary statistics file was finally analyzed in LDSC in one of two ways, as described in the section below.

#### Analysis of Simulated Summary Statistics

We compared the ability to recover the population liability scale heritability 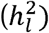 for two approaches: 1) the standard procedure of inputting the total sample prevalence (*v*_*Total*_) and the total sample size (*N*_*Total*_) versus, 2) our proposed approach of inputting the sum of effective sample sizes (∑ *EffN*) and a population prevelance (*P*) of .5 to reflect the fact that the effective sample size equation already accounts for cohort-specific sample ascertainment. For each simulation condition and liability correction approach we report the mean liability scaled heritability estimate, standard deviation across the 100 simulations, the range of parameter estimates, and the mean proportional bias relative to the population generating parameter, calculated as:

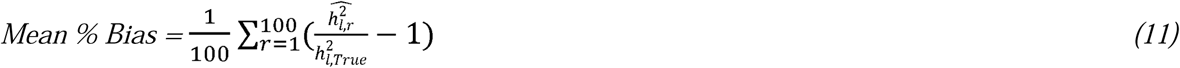

where 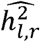 is the parameter estimate for a given run, *r*, and 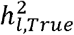 was the population generating value of 15%.

### Analysis of Real Data

We examined liability scale heritability estimates for publicly available, European only summary statistics for 12 major disorders: attention-deficit hyperactivity disorder (ADHD),^20^ alcohol dependence (ALCH)^10^ Alzheimer’s disease (ALZ),^21^ anorexia nervosa (AN),^22^ autism-spectrum disorder (ASD),^9^ bipolar disorer (BIP),^2^ cannabis use disorder (CUD),^23^ major depressive disorder (MDD),^24^ obsessive-compulsive disorder (OCD),^25^ post-traumatic stress disorder (PTSD),^26^ schizophrenia (SCZ),^27^ and Tourette’s syndrome (TS).^28^ For each set of summary statistics, we followed the standard quality control procedure of filtering out SNPs with an imputation quality (INFO) score < .9 and minor allele frequency (MAF) < 1% and filtering to SNPs present in the HapMap3 file excluding the MHC region. In line with prior work for ALZ, we also removed the APOE region prior to calculating heritability. In addition, for ALZ we confirmed that the GERAD consortium was analyzed as a single cohort while the remaining contributing consortiums (ADGC, CHARGE, and EADI) reflected meta-analyzed summary statistics obtained from individual cohorts. Thus, a single ∑ *EffN* was calculated for GERAD while ∑ *EffN* was calculated for each of the contributing cohorts for the other consortiums prior to summing them all together to produced a single ∑ *EffN* for ALZ. For all traits, the liability scale heritability was then calculated using either our proposed approach of inputting ∑ *EffN* or the field standard approach of using *v*_*Total*_. For ∑ *EffN*, the SNP-specific sum of effective sample sizes was used when available. Similarly, when using *v*_*Total*_ the SNP-specific total sample size was utilized when this information was available. Bias was calculated here as the proportion of the ∑ *EffN* estimate captured by *v*_*Total*_ :

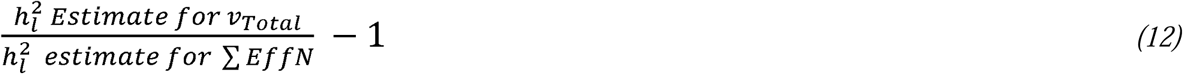

## Data Availability

The data that support the findings of this study are all publicly available.
Summary statistics for data from the Psychiatric Genomics Consortium (PGC) for ADHD, ALCH, AN, ASD, BIP, CUD, MDD, OCD, PTSD, SCZ and TS can be downloaded here:
https://www.med.unc.edu/pgc/download-results/
The summary statistics for ALZ can be found here:
https://www.niagads.org/datasets/ng00075
LD-scores and reference files used to estimate LD-score regression can be downloaded here:
https://alkesgroup.broadinstitute.org/LDSCORE/

https://www.med.unc.edu/pgc/download-results/

https://www.niagads.org/datasets/ng00075

https://alkesgroup.broadinstitute.org/LDSCORE/

## Acknowledgements

ADG was supported by NIH Grants R01MH120219 and RF1AG073593. EMTD was supported by NIH grants R01MH120219 and RF1AG073593 and the Jacobs Foundation. EMTD is a faculty associate of the Population Research Center at the University of Texas, which is supported by NIH grant P2CHD042849. MGN is additionally supported by ZonMW grants 849200011 and 531003014 from The Netherlands Organisation for Health Research and Development, a VENI grant awarded by NWO (VI.Veni.191G.030), NIH grant R01MH120219 and is a Jacobs Foundation Fellow. J.F. is member of the Population Research Center (PRC) and Center on Aging and Population Sciences (CAPS) at The University of Texas at Austin, which are supported by National Institutes of Health (NIH) grants P2CHD042849 and P30AG066614, respectively.

## Notes

### Competing Interest Statement

The authors have declared no competing interest.

### Author Declarations

The primary analyses presented in the paper are for simulation results only and do not use any human subject data. For the real data analyses, we have used de-identified GWAS summary statistics. For those no IRB oversight is necessary. Summary statistics used in the present analyses are publicly available for download.

