## Supplementary Note for "Pervasive Downward Bias in Estimates of Liability Scale Heritability in GWAS Meta-Analysis: A Simple Solution"

**S1. Properties of the Standard GWAS Model**

The standard additive GWAS model estimates the marginal effect size, *b*, from the linear regression phenotype *y* on genetic variant *x*, can be written as:

*y = bx + e*	.

The sampling variance $\sigma_{b}^{2}$ (i.e. the squared standard error, *SE*) of the *b* estimate is given as:

$$\sigma_{b}^{2}={({SE}_{b})}^{2}= \frac{\sigma_{e}^{2}}{\sigma_{x}^{2}n}= \frac{\sigma_{y}^{2}-\sigma_{x}^{2}(b^{2})}{\sigma_{x}^{2}n} ,$$

where $\sigma_{e}^{2}$ is the residual variance of *y*, *n* is the sample size, $\sigma_{y}^{2}$ is the variance of *y*, and $\sigma_{x}^{2}$ is the variance of the variant. Without loss of generality, we assume that *y* and *x* have each been standardized to mean 0, and standard deviation 1, such that $\sigma_{b}^{2}$ reduces to:

$$\sigma_{b}^{2}={({SE}_{b})}^{2}= \frac{\sigma_{e}^{2}}{n}= \frac{1-b^{2}}{n} .$$

Because GWAS effect sizes for complex traits are extremely small, $1-b^{2}\approx1$ such that:

$$\sigma_{b}^{2}={({SE}_{b})}^{2}\approx\frac{1}{n} ,$$

$$Z=\frac{b}{SE}=\frac{b}{\frac{1}{\sqrt{n}}}=b\sqrt{n} ,$$

$$Z^{2}=\chi^{2}={nb}^{2} .$$

**S2. Estimating Heritability and Genetic Covariance from GWAS Summary Statistics**

We can model *K* phenotypes and *M* SNPs measured in *N* individuals according to the equation:

$$y_{i,k}=x_{i,j}\beta_{j,k}+\epsilon_{i,k}$$

where $y_{i,k}$is the score for person *i* on the standardized phenotype *k,* $x_{i,j}$is the standardized genotype for person *i* on SNP *j,* $\beta_{j,k}$ is the true standardized effect size for SNP *j* on phenotype *k,* and $\epsilon_{i,k}$ is the residual for person *i* on phenotype *k*. This model can be written in matrix form as:

$$Y=XB+E$$

where *Y* is an *N× K* matrix of standardized scores for person *i* on phenotype *k, X* is an *N×M* matrix of standardized genotypes for person *i* on SNP *j, B* is an *M×K* matrix of true standardized genotype effect sizes for SNP *j* on phenotype *k,* and *E* is an *N×K* matrix of residuals for person *i* on phenotype *k*.

Assuming independence of genotypes $x_{i,j}$ from one another, but incorporating each genotype’s average linkage disequilibrium with the others for model estimation, LD Score regression^1–3^ (LDSC) is used to model *β_j,k_* as phenotype-specific random effects, varying over SNPs, with $\mathbb{E}\left[ B \right]=0$ and $cov\left[ B \right]=\frac{1}{M}S$. The diagonal elements of *S* contain the SNP heritability ($h_{SNP}^{2})$, and the off-diagonal elements of *S* contain the genetic covariances ($\sigma_{g}$) between phenotypes, i.e. the genetic correlations between phenotypes scaled relative to the SNP heritabilities of the respective phenotypes. The elements of *S* are estimated from GWAS summary statistics by regressing the product of *Z* statistics for the linear regression of phenotypes 1 and 2 on SNP *j* on the LD score of SNP *j* and solving for $\sigma_{g}$ as follows:

$$\mathbb{E}\left[ Z_{1j}Z_{2j} \right]=\sqrt{N_{1}N_{2}}\frac{\sigma_{g}}{M}\mathcal{l}\left( j \right)+\frac{\rho N_{s}}{\sqrt{N_{1}N_{2}}}+a ,$$

where *N_1_* and *N_2_* are the sample sizes for phenotypes 1 and phenotypes 2, *M* is the number of SNPs, *ℓ*(*j*) is the LD score of SNP *j* (that is, the sum of squared correlations between SNP *j* and all other SNPs in the reference panel), *N_s_* is the number of individuals included in both GWAS samples, *ρ* is the phenotypic correlation within the overlapping samples, and *a* is a term representing unmeasured sources of confounding such as shared population stratification across GWASs. When the *Z* statistics for the same phenotype are double entered into the left-hand side of the above equation, such that $\mathbb{E}\left[ Z_{1j}Z \right]$ becomes $\mathbb{E}\left[ Z_{j}^{2} \right]=\mathbb{E}\left[ \chi_{j}^{2} \right]$, the equation reduces to the univariate LDSC model, and $\sigma_{g}$ becomes an estimate of $h_{SNP}^{2}$, as follows

$$\mathbb{E}\left[ Z_{j}^{2} \right]=\mathbb{E}\left[ \chi_{j}^{2} \right]=N\frac{h_{SNP}^{2}}{M}\mathcal{l}\left( j \right)+a+1 .$$

Without loss of generality, we focus here on the univariate LDSC model.

The $h_{SNP}^{2}$ estimate in LDSC is determined from the LDSC Slope via the coefficient $N\frac{h_{SNP}^{2}}{M}$. Noting that $\frac{h_{SNP}^{2}}{M}$ is the average variance explained per SNP, it follows that:

$$LDSC Slope=N var\left( \beta_{j,k} \right)=N\bar{\beta_{j,k}^{2}}$$

Noting further that LD Slope relates the LD score to the squared Z statistic, $\mathbb{E}\left[ Z_{j}^{2} \right]$, we can see that the foundation of this association is the simple relation between Z and the standardized linear regression coefficient, derived earlier as:

$$Z^{2}={nb}^{2} .$$

For GWAS of binary traits, the heritability estimate from the standard LDSC equation produces an estimate of “observed scale” SNP heritability that depends on the sample prevalence, which for ascertained samples, will not correspond to the population prevalence and may vary considerably across studies. This observed scale SNP heritability can be transformed into a more interpretable liability scale SNP heritability estimate; i.e. the heritability of the continuous underlying liability toward the binary outcome that does not depend on sample prevalence. Liability scale heritability ${(h}_{l}^{2})$ can be computed from observed scale heritability ${(h}_{o}^{2})$ as:^4–6^

$${h_{l}^{2}=h}_{o}^{2}\frac{P^{2}\left( 1-P \right)^{2}}{\phi^{2}v\left( 1-v \right)} ,$$

or more directly computed in the reduced form LDSC equation for binary traits as follows:

$$\mathbb{E}\left[ \chi_{j}^{2} \right]=\frac{\phi^{2}}{P^{2}\left( 1-P \right)^{2}}v\left( 1-v \right)N \frac{h_{l}^{2}}{M}\mathcal{l}\left( j \right)+a+1 ,$$

where $v$ is the sample prevalence, *P* is the population prevalence, and $\phi$ is the height of the standard normal density of at the threshold corresponding to *P.* We compare the LDSC slope to the $\chi^{2}$ statistic for the transformed regression coefficient appropriate for binary traits in section **S4**.

**S3. GWAS Meta-Analysis of Quantitative Traits**

An inverse variance weighted meta-analysis can be used to combine effect size estimates, $b_{k}$, across *k* independent sets of GWAS summary statistics using weights $w_{k}=\frac{1}{\sigma_{b_{k}}^{2}}$. (de la Fuente et al.^7^ formally demonstrate the equivalence of inverse-variance weighted meta-analysis of effect sizes and sample-size weighted meta-analysis of Z statistics.) For continuously distributed phenotypes (quantitative traits), effect sizes from linear regression are appropriate to combine directly. In this circumstance, $\frac{1}{\sigma_{b_{k}}^{2}}\approx n$ such that:

$$b_{inv var}=\frac{\sum w_{k}b_{k}}{\sum w_{k}}= \frac{\sum\frac{1}{\sigma_{b_{k}}^{2}}b_{k}}{\sum\frac{1}{\sigma_{b_{k}}^{2}}}=\frac{\sum n_{k}b_{k}}{\sum n_{k}} ,$$

$${SE}_{b_{inv var}}=\frac{1}{\sqrt{\sum w_{k}}}= \frac{1}{\sqrt{\sum\frac{1}{\sigma_{b_{k}}^{2}}}}=\frac{1}{\sqrt{\sum n_{k}}} ,$$

$$Z_{inv var}= \frac{b_{inv var}}{{SE}_{b_{inv var}}}=\frac{b_{inv var}}{\frac{1}{\sqrt{\sum n_{k}}}}= b_{inv var}\sqrt{\sum n_{k}} ,$$

where $b_{inv var}$ is the fixed effects meta-analytic estimate of *b*. It follows that:

$$\chi_{inv var}^{2}=Z_{inv var}^{2}= b_{inv var}^{2}\sum n_{k} ,$$

which can be compared to the form of the LDSC slope for continuous traits:

$$LDSC Slope=\bar{\beta_{j,k}^{2}} N ,$$

thus indicating that when using meta-analytic summary statistics for traits analyzed via linear GWAS, $\sum n_{k}$ (i.e., the total sample size) is appropriate to enter for *N* in LDSC, as is standard practice.

**S4. GWAS Meta-Analysis of Binary Traits**

When the GWAS phenotype is a binary trait, the effect sizes, $b_{k}$, from linear regression must be transformed such that they are comparable metrics across GWAS before they can be combined via meta-analysis. This is of particular relevance for ascertained samples (where cases are typically oversampled) in which the degree of ascertainment varies across contributing GWAS. This is because, under a standard polygenic model of disease liability, the linear slope relating the allele count to the binary phenotype (the slope of the so-called linear probability function) depends on the sample prevalence. In contrast, because odds ratios are not dependent on sample prevalence, the logistic regression coefficient is an appropriate effect for placing GWAS on the same metric. In the context of GWAS of complex traits where individual SNP effects are extremely small, the regression coefficient for the logistic regression of the binary phenotype on the standardized variant ($b_{logit STDX}$) can be closely approximated from the coefficient from the linear regression of the standardized binary phenotype on the standardized variant *X* ($b_{linear STD}$) as:^8,9^

$$b_{logit STDX}\approx\frac{b_{linear STD}}{\sqrt{v\left( 1-v \right)}} ,$$

$$\sigma_{b_{logit STDX}}^{2}={({SE}_{b_{logit STDX}})}^{2}\approx\frac{\sigma_{b_{linear STD}}^{2}}{v\left( 1-v \right)}= \frac{1}{v\left( 1-v \right)n} ,$$

$$Z=\frac{b_{logit STDX}}{{SE}_{b_{logit STDX}}}=\frac{b_{logit STDX}}{\frac{1}{\sqrt{v\left( 1-v \right)n}}}=b_{logit STDX}\sqrt{v\left( 1-v \right)n} ,$$

where *v* is the proportion of cases, such that *v(1-v)* is the observed variance of the binary phenotype.

We use this relation to derive a corrected linear effect size, $b^{*}$, that would have been obtained had the case-control GWAS been on a balanced sample (i.e. 50% cases, 50% controls). We compute $b^{*}$ by substituting .5 for *v* into the equation approximating logistic regression from linear regression:

$$b_{logit STDX}\approx\frac{b_{linear STD}}{\sqrt{v\left( 1-v \right)}} =\frac{b^{*}}{\sqrt{.5\left( 1-.5 \right)}} ,$$

$$\sqrt{.5\left( 1-.5 \right)}\frac{b_{linear STD}}{\sqrt{v\left( 1-v \right)}} =b^{*} ,$$

$$b^{*}=.5\frac{b_{linear STD}}{\sqrt{v\left( 1-v \right)}} ,$$

$$Z=b_{logit STDX}\sqrt{v\left( 1-v \right)n}=\frac{b^{*}}{\sqrt{.5\left( 1-.5 \right)}}\sqrt{v\left( 1-v \right)n} = 2b^{*}\sqrt{v\left( 1-v \right)n} ,$$

therefore,

$$Z^{2}= {{(b}^{*})}^{2}v\left( 1-v \right)4n .$$

The SE of $b^{*}$ is derived as:

$Z=2b^{*}\sqrt{v\left( 1-v \right)n}=\frac{b^{*}}{{SE}_{b*}}$,

${SE}_{b*}=\frac{1}{2\sqrt{v\left( 1-v \right)n}}$ ,

$\sigma_{b*}^{2}={({SE}_{b*})}^{2}=\frac{1}{4v\left( 1-v \right)n}$ .

Noting that the standard term for SE in OLS is $\frac{1}{n}$, it is sensible to refer to $4v\left( 1-v \right)n$ as the *effective sample size (EffN)* for a balanced design.

Because $b^{*}$ represents the effect size that would have been obtained had all studies had balanced ascertainment, they are on the same scale as one another and can be meta-analyzed. We can specify the inverse weighted meta-analysis of $b^{*}$, its *SE*, and its *Z* and $\chi^{2}$ statics as:

$$b_{inv var}^{*}=\frac{\sum w_{k}b_{k}^{*}}{\sum w_{k}}=\frac{\sum{{4v}_{k}(1-v_{k})n}_{k}b_{k}^{*}}{\sum{{4v}_{k}(1-v_{k})n}_{k}} =\frac{\sum{{4v}_{k}(1-v_{k})n}_{k}.5\frac{b_{linear STD}}{\sqrt{v\left( 1-v \right)}}}{\sum{{4v}_{k}(1-v_{k})n}_{k}}= \frac{\sum{\sqrt{v\left( 1-v \right)}n}_{k}b_{linear STD}}{2\sum{v_{k}(1-v_{k})n}_{k}},$$

$${SE}_{b_{inv var}^{*}}=\frac{1}{\sqrt{\sum w_{k}}}= \frac{1}{\sqrt{\sum{{4v}_{k}(1-v_{k})n}_{k}}} ,$$

$$Z_{inv var}^{*}= \frac{b_{inv var}^{*}}{{SE}_{b_{inv var}^{*}}}=\frac{b_{inv var}^{*}}{\frac{1}{\sqrt{\sum4{v_{k}(1-v_{k})n}_{k}}}}= b_{inv var}^{*}\sqrt{\sum{{4v}_{k}(1-v_{k})n}_{k}} ,$$

where $b_{inv var}^{*}$ is the fixed effects meta-analytic estimate of $b^{*}$. Note that, apart from a difference in scaling constants, this is equivalent to an inverse-variance weighted meta-analysis of the logistic regression coefficients themselves.

We can see that the relation between squared *Z* statistics and the squared meta-analytic effect size is governed by:

$${\chi^{*}}_{inv var}^{2}={Z^{*}}_{inv var}^{2}= {b^{*}}_{inv var}^{2}\sum{{4v}_{k}\left( 1-v_{k} \right)n}_{k}$$

$$={b^{*}}_{inv var}^{2}\sum{EffN}_{k} ,$$

which can be compared to the form of the LDSC slope

$$LDSC Slope=\bar{\beta_{j,k}^{2}} N ,$$

thus indicating that $\sum EffN$ is appropriate to use for LDSC of binary traits to produce an unbiased estimate of observed scale heritability for a balanced design, $h_{o^{*}}^{2}$. Liability scale heritability can then be obtained from the following, which now only includes terms that are constant across samples:

$${h_{l}^{2}=h}_{o^{*}}^{2}\frac{P^{2}\left( 1-P \right)^{2}}{\phi^{2}.5(1-.5)}$$

Liability scale heritability can similarly be derived for the reduced form equation by substituting $\sum EffN$ for *N* and setting $v$ to .5 to reflect the fact that *EffN* corresponds to a balanced design, yielding the following for the LDSC slope:

$$LDSC Slope=\frac{\phi^{2}}{P^{2}\left( 1-P \right)^{2}}v\left( 1-v \right)\left( \sum{EffN}_{k} \right)\frac{h_{l}^{2}}{M} = \frac{\phi^{2}}{P^{2}\left( 1-P \right)^{2}}.5\left( 1-.5 \right)\frac{h_{l}^{2}}{M}\sum{4v_{k}\left( 1-v_{k} \right)n}_{k}=\frac{\phi^{2}}{P^{2}\left( 1-P \right)^{2}}\frac{h_{l}^{2}}{M}\left( \sum{v_{k}\left( 1-v_{k} \right)n}_{k} \right) .$$

Of particular relevance here is that the term in the LDSC equation that involves the product of sample size, case proportions, and control proportions must be derived via an *n*-weighted summation of individual case proportions for each set of summary statistics in the meta-analysis, not as the product of total sample size and the proportion of total cases and controls in the full, meta-analytic sample. In other words:

$$\sum{v_{k}\left( 1-v_{k} \right)n}_{k}\neq\frac{\left( \sum{v_{k} n}_{k} \right)}{\sum n_{k}}\frac{\left( \sum{{(1-v}_{k}) n}_{k} \right)}{\sum n_{k}}\left( \sum n_{k} \right)=\left( \frac{N_{Cases Total}}{N_{Total}} \right)\left( \frac{N_{Controls Total}}{N_{Total}} \right)N_{Total},$$

or similarly put:

$$\sum EffN=\sum{{4v}_{k}\left( 1-v_{k} \right)n}_{k}\neq4\frac{\left( \sum{v_{k} n}_{k} \right)}{\sum n_{k}}\frac{\left( \sum{{(1-v}_{k}) n}_{k} \right)}{\sum n_{k}}\left( \sum n_{k} \right)=4\left( \frac{N_{Cases Total}}{N_{Total}} \right)\left( \frac{N_{Controls Total}}{N_{Total}} \right)N_{Total}.$$

The equations on the left of the inequalities represent the correct terms, whereas the equations on right of the inequalities represent how the sample size and proportion of cases in meta-analysis are conventionally entered into LDSC when calculating $h_{l}^{2}$ from GWAS meta-analysis of binary phenotypes. In other words, the inequality indicates that the conventional approaches are incorrect.
